# An ECG foundation model for generalizable cardiac function prediction across the lifespan

**DOI:** 10.64898/2026.05.26.26354128

**Authors:** Yuting Yang, Lorenzo Peracchio, Joshua Mayourian, Timothy Miller, William G. La Cava

## Abstract

**Background:** Artificial intelligence-enhanced electrocardiography (AI-ECG) enables scalable, low-cost cardiac dysfunction screening, but existing models are annotation-intensive and predominantly adult-derived, leaving paediatric generalizability uncertain. Paediatric cohorts exhibit highly variable cardiac morphology and function compared to adults, which may be useful for learning generalizable AI-ECG models.

**Methods:** We pretrained ECG-Fyler on a predominantly paediatric, all-age cohort at Boston Children’s Hospital (1992–2023), annotated with a cardiology-specific coding system (Fyler codes), and evaluated it on assessments from echocardiography (echo) and cardiac magnetic resonance (CMR) studies. We validated on an external adult cohort from Columbia University Irving Medical Center. Performance was benchmarked against several AI-ECG foundation models by AUROC across age groups, lesion types, and limited-data scenarios.

**Findings:** The pretraining cohort comprised 782,138 ECGs from 255,271 patients (median age: 10.9 years, IQR: [2.8–16.8]). Internal evaluation included 178,495 ECG-echo pairs (median age: 10.9 [3.7–17.0]) and 8,584 ECG-CMR pairs (median age: 20.7 [15.6–29.6]). External validation included 82,543 ECG-echo pairs from adults (median age: 64.0 [52.0–74.0]). ECG-Fyler improved AUROC across biventricular dysfunction and dilation tasks, with the largest gains in low-data settings. In internal validation, ECG-Fyler detected low left ventricular ejection fraction (LVEF***≤***40%) from only 100 fine-tuning samples (AUROC: 0.80, 95% CI: [0.78–0.80]), outperforming other models (AUROC ***<***0.65) and improving with additional fine-tuning (AUROC: 0.94 [0.93–0.94]). Similar improvements were observed for CMR-derived LVEF, RVEF, and ventricular dilation. In external validation on adults, ECG-Fyler exhibited an AUROC of 0.83 (CI: [0.82–0.85]) for LVEF***≤***40%. After fine-tuning on less than 10% of external data, LVEF ***≤***45% performance (AUROC: 0.87 [0.86–0.88]) outperformed a fully trained, site-specific prior model (AUROC: 0.85 [0.84–0.87]).

**Interpretation:** Pretraining on richly annotated, paediatric-dominant ECGs yields models that transfer efficiently across institutions and ages, supporting AI-ECG screening and triage when labels or imaging access are limited.

**Funding:** National Institutes of Health (R01LM012973); Kostin Innovation Fund, Boston Children’s Hospital.

**Research in context:** *Evidence before this study:* On April 29, 2026, we searched PubMed from database inception to April 29, 2026, without language restrictions, using the terms “electrocardiogram” AND “foundation model”. We considered original studies describing ECG foundation models, transferable representation- learning approaches, or mixed-age AI-ECG studies relevant to cardiac function prediction, and excluded non-ECG studies, non-original research, and purely task-specific models without a transferable pretraining component. The available evidence consisted mainly of heterogeneous observational development and validation studies, so we did not do a formal meta-analysis. We identified 11 records. Most published artificial intelligence-enhanced electrocardiogram (AI-ECG) foundation models were developed in adult cohorts, whereas paediatric evaluation remained limited. We did not identify a previous report describing a single ECG foundation model trained on a paediatric-dominant, all-age cohort and evaluated across paediatric and adult populations using both echocardiography- and cardiac magnetic resonance-derived measures of ventricular function.

*Added value of this study:* We developed ECG-Fyler, a clinically grounded ECG foundation model pretrained on 782,138 ECGs from 255,271 patients in a paediatric-dominant, all-age cohort using structured Fyler code annotations. We evaluated transfer learning across echocardiography- and cardiac magnetic resonance-derived tasks, congenital heart disease lesion subgroups, low-resource fine-tuning scenarios, and external adult validation. ECG-Fyler consistently outperformed training from scratch and other ECG foundation-model baselines, with the largest gains when labelled data were scarce, and showed strong cross-age and cross-institution generalization from paediatric pretraining to adult external validation.

*Implications of all the available evidence:* Taken together, the available evidence suggests that ECG foundation models can improve data efficiency and generalizability, but the field remains dominated by adult data and task-specific applications. Our findings extend this evidence by suggesting that clinically grounded supervised pretraining on a paediatric-dominant, lifespan-spanning ECG corpus can support generalizable prediction of ventricular dysfunction and dilation across age groups and institutions. If validated prospectively, such models could support lower-cost screening, triage, and longitudinal monitoring to help prioritize downstream echocardiography or cardiac magnetic resonance imaging, particularly in congenital heart disease and other settings where labelled imaging data are limited.

## 1 Introduction

An electrocardiogram (ECG) is a noninvasive recording of cardiac electrical activity obtained from body-surface voltage measurements. As a rapid, standardized, and costeffective test, ECG is widely used for cardiovascular diagnosis and initial screening^[1]^. Recent advances in artificial intelligence–enhanced ECG (AI-ECG) have extended its utility beyond waveform interpretation to the prediction of complex structural and functional cardiac phenotypes. Phenotypes include left ventricular ejection fraction (LVEF) ^[2–4]^, ventricular dilation^[5]^, and tricuspid regurgitation^[6]^, all of which traditionally require resource-intensive imaging modalities such as echocardiography (echo) and cardiac magnetic resonance imaging (CMR). In this context, AI-ECG provides a scalable and cost-effective strategy to identify high-risk individuals and guide downstream imaging, facilitating earlier detection and more efficient clinical triage. These advances have progressed toward real-world clinical deployment: an AI-ECG algorithm for detecting reduced ejection fraction has been validated in prospective, cluster-randomised trials and received FDA clearance^[2,7]^, while broad AI-ECG detection of structural heart disease has recently been evaluated in a prospective clinical trial^[8]^.

Despite these advances, most existing AI-ECG models remain confined to specific diagnostic tasks and datasets. Their ability to generalize across cohorts from different institutions, age groups, or congenital heart disease (CHD) subtypes remains incompletely characterized. The practical challenge of training ECG models from scratch or adapting them to small care centers with limited data and resources limits their real-world applicability. In this light, there is a clear need for general and transferable AI-ECG tools.

Foundation models provide a promising solution to this general problem by creating reusable, task-agnostic data representations that can be adapted across diverse clinical settings. Existing ECG foundation models can be broadly categorized into two approaches: *self-supervised* learning on large, un-annotated ECG datasets, and *supervised* learning which levarages ECG-linked clinical reports or diagnostic annotations. Representative self-supervised approaches, such as ECG-FM^[9]^, are trained to reconstruct masked portions of ECG signals and identify portions of signal from the same reading, as a proxy for learning generalizable ECG representations. In contrast, supervised approaches like ECGFounder^[10]^ are trained to estimate widely available values pulled from clinical reports, and then transferred to real-world tasks. Related work has also explored masked modeling of ECG time–frequency representations and transformer-based architectures pretrained on large inpatient ECG cohorts^[11,12]^. These models exhibit strong and scalable performance on downstream tasks, yet the vast majority are developed and evaluated on adult populations; systematic evaluation of their generalization to paediatric cohorts remains limited. Furthermore, no existing work has considered the development of ECG foundation models from predominantly paediatric cohorts, which, due to their representation of developing hearts, may capture distinct ECG patterns and phenotypic heterogeneity that subsume and expand upon the ECGs represented in adult cohorts.

To address these limitations, we developed ECG-Fyler, a clinically grounded ECG foundation model trained on 782,138 ECGs annotated with the Fyler Code System, a hierarchical and fine-grained coding system that provides standardized descriptions of diagnoses, findings, and interventions^[13]^. Compared with free-text–derived labels and other coding systems, Fyler codes offer a structured and information-rich annotation ontology, enabling dense and clinically consistent supervision for representation learning. Importantly, our dataset is paediatric-dominant but spans the full age spectrum, including infants, children, adolescents, and adults, thereby capturing heterogeneous ECG patterns from developing hearts while maintaining strong representation of both paediatric and adult populations, including CHD patients. The resulting dataset provides a uniquely structured and developmentally diverse resource for learning clinically meaningful ECG representations, with the goal of broad transferability to new populations.

ECG-Fyler is a ResNet-based foundation model^[14]^, pretrained to predict 93 Fyler Code labels, producing ECG embeddings that encode rich structural and functional cardiac information. These representations are subsequently evaluated across multiple downstream tasks derived from echo and CMR, including LVEF, ventricular dilation, and right ventricular functional assessment across both congenital and non-congenital cardiac populations. We show that the resulting ECG representations improve prediction of clinically relevant cardiac measurements from both echo and CMR, generalizing robustly across institutions, age groups, and CHD categories. ECG-Fyler consistently outperforms existing ECG foundation models on paediatric and adult cohorts. These performance gains are particularly robust under limited-data settings (e.g., limiting training to 100 labeled samples), highlighting strong transferability to low-resource clinical settings. As a result, ECG-Fyler has the potential to improve access to AI-ECG by performing well with less site-specific data compared to prior approaches. Such models support several clinical use cases, including early screening of cardiac function in populations with constrained access to advanced imaging, longitudinal monitoring of structural progression in patients with known lesions, and triage of patients requiring prioritization for echo or CMR evaluation.

## 2 Methods

### 2.1 Study population

We collected patient data from Boston Children’s Hospital (BCH) spanning 1992 to January 2023. ECG records were linked to the institutional Fyler Coding System^[13]^, a hierarchical clinical ontology for congenital heart disease comprising more than 3,000 structured labels covering ECG findings, echo measurements, surgical procedures, and catheter-based interventions. We excluded ECGs associated only with rare Fyler Codes, defined as codes appearing in fewer than 10 ECGs in the dataset, to ensure sufficient label support for model pretraining. We retained only ECG-related Fyler Codes (93 codes), capturing common conduction abnormalities, repolarization changes, ventricular hypertrophy, axis deviations, and rhythm disturbances. This restriction focuses representation learning on ECG signals and reduces potential information leakage from non-ECG modalities, such as echo-related annotations, during the pretraining stage.

Validation was performed on internal and external cohorts. The internal validation cohort comprised patients from the same paediatric hospital, but excluded from the pretraining dataset. The external validation cohort was a publicly available adult dataset^[15]^ from Columbia University Irving Medical Center (CUIMC). To avoid data leakage, all patients included in downstream task test sets were excluded from the pretraining cohort.

We evaluated ECG-Fyler on two downstream tasks. The first involved echo-based measurements to assess cardiac function from ultrasound images, and the second relied on CMR as a reference standard for structural and functional assessment. The internal echo cohort included patients with and without congenital heart disease (CHD), with lesions classified according to standard clinical definitions^[16]^. CMR imaging has an established role in the lifelong management of patients with CHD, providing accurate assessment of left and right ventricular (LV and RV) size and function, as well as functional single ventricle measurements, which are often challenging to quantify by echo. For CMR analysis, we used patient data from BCH (2002–2021), following previously described cohort construction and preprocessing protocols^[5]^.

### 2.2 Quality control and data preprocessing

ECG waveforms were obtained from the institutional MUSE system (GE Healthcare) as 10-second 12-lead recordings. ECGs shorter than 10 s or missing lead information were discarded. Remaining recordings underwent uniform preprocessing and quality control before model training, including filtering and fixed-length signal trimming; fewer than 2% of ECGs failed quality control. Detailed signal-processing steps are provided in the code repository.

### 2.3 Outcomes

For echo analysis, the primary outcome was LVEF ≤40%, with secondary outcomes including LVEF ≤50% and ≤30%. For CMR, the individual outcomes were greater than mild LVEF (LVEF ≤40%), RV systolic dysfunction (RVEF ≤35%), LV dilation (left ventricular end-diastolic volume [LVEDV] z-score ≥ 4, corresponding to 121 mL/m^2^ in women and 141 mL/m^2^ in men), and RV dilation (RV end-diastolic volume [RVEDV] z-score ≥ 4, corresponding to 130 mL/m^2^ in women and 143 mL/m^2^ in men).

### 2.4 Model selection, architecture, and training

ECG-Fyler was developed from a ResNet-based encoder^[14]^, a widely adopted architecture for AI-ECG^[17–20]^. We compared ECG-Fyler to an identical ResNet trained from scratch (without pretraining) to evaluate the impact of supervised pretraining on Fyler code prediction. Models were trained with the Adam optimizer^[21]^ and early stopping. Additional implementation and hyperparameter details are provided in the code repository.

### 2.5 Baseline models

We evaluated ECG-Fyler against two existing ECG foundation models and one task-specific model. ViTMAE^[22–24]^ served as a self-supervised baseline and was pretrained on the same ECG cohort as ECG-Fyler. ECGFounder^[10]^ served as a large supervised ECG foundation model pretrained on a predominantly adult cohort. We used the released checkpoints for both baselines and fine-tuned or adapted them for the downstream tasks using the same train-test splits as ECG-Fyler. We also report results from EchoMini^[15,25]^, a task-specific model released with the external cohort study, trained at the same site, and reportedfor LVEF ≤ 45% only. Full architectural and implementation details for all comparators are provided in the code repository.

### 2.6 Performance evaluation and statistical analyses

We computed both the area under the receiver operating characteristic curve (AUROC) and the area under the precision-recall curve (AUPRC) to evaluate model performance. Confidence intervals were obtained via resampling with 1,000 bootstraps.

### 2.7 Low-resource training scenarios

To simulate real-world low-resource scenarios with limited annotated training data, we created subsets of the training data by randomly sampling 100 examples, 1%, 10%, and 100% of the full training set. These subsets represent progressively larger training data regimes, enabling evaluation of model performance across different data resource constraints.

### 2.8 Visual Analysis of ECG Representations Across Cohorts

Our ECG representations are trained on a cohort spanning multiple age groups. This mixed age distribution raises the question of whether the learned representations (i.e., the foundation model embeddings) capture generalizable physiological structure beyond paediatric populations.

To investigate this, and to evaluate cross-institutional and cross-age generalization, we extracted ECG-Fyler embeddings for two independent test cohorts (BCH and CUIMC). The external CUIMC cohort originates from a different institution and consists exclusively of adults (*>*18 years) with a substantially older age distribution. Embeddings were obtained from the penultimate layer of the model and visualized using Uniform Manifold Approximation and Projection (UMAP)^[26]^, fitted on BCH embeddings with CUIMC samples projected into the same space. To mitigate potential bias arising from unequal cohort sizes, we randomly subsampled the test embeddings to the same sample size for visualization.

## 3 Results

### 3.1 Pretraining cohort characteristics

The pretraining cohort comprised 782,138 ECG recordings from 255,271 patients (Table 1). Although primarily paediatric, it spanned the full age spectrum (min age: less than 1 year, max age: 84 years; 80.3% younger than 18 years, 19.7% older than 18 years). Fyler-code frequencies ranged from common normal and conduction-pattern labels to rare rhythm and conduction abnormalities, reflecting broad electrophysiologic heterogeneity. The full code distribution is provided in the Supplementary Tables.

**Table 1.** Composition of the all-age pretraining ECG cohort. Summary of dataset size, demographics, and Fyler code distribution.

|  |  |
| --- | --- |
| ECGs with Fyler Codes | 782,138 |
| Patients | 255,271 |
| Female | 128,135 (50.2%) |
| Age at ECG, years | 10.9 (IQR 2.8–16.8) |
| Paediatric (0-18) | 630,846 (80.7%) |
| Adult ($\geq 18$) | 151,292 (19.3%) |
| Fyler codes (highest-frequency) |  |
| Normal ECG | 332,708 (42.5%) |
| Incomplete right bundle branch block (RSR') | 101,333 (13.0%) |
| ST-T wave change, non-specific | 80,560 (10.3%) |
| Right bundle branch block (complete) | 67,800 (8.7%) |
| Right ventricular hypertrophy (ECG) | 60,619 (7.8%) |
| Axis deviation, right (ECG) | 59,606 (7.6%) |
| Axis deviation, superior (ECG) | 36,500 (4.7%) |
| QTc prolonged | 35,974 (4.6%) |
| Sinus tachycardia | 35,971 (4.6%) |
| Sinus arrhythmia | 34,005 (4.3%) |
| Fyler codes (lowest-frequency) |  |
| ECG Digoxin effect | 70 (0.009%) |
| Multifocal atrial tachycardia | 67 (0.009%) |
| Mahaim fiber | 65 (0.008%) |
| Left posterior hemiblock | 59 (0.008%) |
| Junctional rhythm nodal | 39 (0.005%) |
| Complete heart block (Acquired) | 32 (0.004%) |
| Pulmonary vein abnormality | 22 (0.003%) |
| Infarct, anteroseptal | 16 (0.002%) |
| Supraventricular tachycardia | 16 (0.002%) |
| Supraventriculr tachycardia wide QRS | 10 (0.001%) |

Data are reported as n or n (%). All Fyler codes and age strata statistics are based on ECG-level observations.

### 3.2 Evaluation cohort characteristics

We evaluated ECG-Fyler on internal BCH echo and CMR cohorts and an external adult echo cohort from CUIMC (Table 2). The internal test sets had no patient overlap with the pretraining or fine-tuning cohorts. The internal echo cohort contained 124,265 training and 54,230 testing ECG–echo pairs from predominantly paediatric patients, whereas the internal CMR cohort contained 6,833 training and 1,751 testing ECG–CMR pairs and enabled assessment of biventricular function and dilation.

**Table 2.** Characteristics of the evaluation cohorts. Summary of internal (BCH) and external (CUIMC) cohorts, including sample sizes, demographics, and clinical outcomes across training and testing splits.

|  | Internal (BCH) |  | External (CUIMC) |  |
| --- | --- | --- | --- | --- |
|  | Training | Testing | Training | Testing |
| **Echo** |  |  |  |  |
| ECG-Echo pairs | 124,265 | 54,230 | 77,101 | 5,442 |
| Patients | 49,158 | 21,068 | 30,844 | 5,442 |
| Age at ECG, years | 10.5 (3.5-16.8) | 10.9 (3.7-17.0) | 63.0 (52.0-73.0) | 64.0 (52.0-74.0) |
| Female at ECG (%) | 22,835 (46.5%) | 9,813 (46.6%) | 33,524 (46.3%) | 2,731 (50.2%) |
| Measurements |  |  |  |  |
| LVEF ≤ 50% | 8,525 (6.9%) | 3,674 (6.8%) | 21,321 (27.7%) | 1,185 (21.8%) |
| LVEF ≤ 40% | 3,381 (2.7%) | 1,473 (2.7%) | 14,902 (19.3%) | 761 (14.0%) |
| LVEF ≤ 30% | 1,490 (1.2%) | 598 (1.1%) | 9,820 (12.7%) | 463 (8.5%) |
| **CMR** |  |  |  |  |
| ECG-CMR pairs | 6,833 | 1,751 | - | - |
| Patients | 3,954 | 987 | - | - |
| Age at ECG, years | 20.7 (15.5-30.4) | 20.7 (15.6-29.6) | - | - |
| Female at ECG (%) | 2,972 (43.5%) | 747 (42.7%) | - | - |
| Measurements |  |  |  |  |
| LVEF ≤ 40% | 231 (3.4%) | 54 (3.1%) | - | - |
| RVEF ≤ 35% | 300 (4.4%) | 85 (4.9%) | - | - |
| LVEDV z-score ≥ 4 | 566 (8.3%) | 112 (6.4%) | - | - |
| RVEDV z-score ≥ 4 | 1,246 (18.2%) | 341 (19.5%) | - | - |

Data are n, n (%), or median (IQR).

The external CUIMC cohort provided a marked age shift (median age: 64.0, IQR: [52.0–74.0]) and disease-spectrum shift relative to BCH, including a higher prevalence of reduced LVEF (Table 2). The BCH cohort also captured heterogeneous CHD anatomy, with similar lesion distributions across training and testing splits (Table 3), enabling subgroup analyses across common and rare lesion categories.

**Table 3.** Distribution of congenital heart disease lesions in the BCH cohort.

|  | Training | Testing |
| --- | --- | --- |
| Ventricular septal defect | 21,171 (17.0%) | 8,926 (16.5%) |
| Cardiomyopathy | 18,509 (14.9%) | 8,082 (14.9%) |
| Atrial septal defect | 14,860 (12.0%) | 6,171 (11.4%) |
| Coarctation of the aorta | 12,353 (9.9%) | 4,998 (9.2%) |
| Tetralogy of fallot | 8,980 (7.2%) | 4,108 (7.6%) |
| D-loop TGA | 4,313 (3.5%) | 1,982 (3.7%) |
| Hypoplastic left heart syndrome | 4,050 (3.3%) | 1,689 (3.1%) |
| Pulmonary atresia | 3,994 (3.2%) | 1,808 (3.3%) |
| Pacemaker | 2,732 (2.2%) | 1,145 (2.1%) |
| Double outlet right ventricular | 2,720 (2.2%) | 1,042 (1.9%) |
| Complete atrioventricular canal | 2,640 (2.1%) | 1,130 (2.1%) |
| Ebstein | 1,481 (1.2%) | 676 (1.2%) |
| Total anomalous pulmonary venous return | 1,463 (1.2%) | 758 (1.4%) |
| Truncus arteriosus | 1,461 (1.2%) | 548 (1.0%) |
| Dextrocardia | 1,265 (1.0%) | 403 (0.7%) |
| L-loop TGA | 1,191 (1.0%) | 399 (0.7%) |
| Tricuspid atresia | 1,117 (0.9%) | 358 (0.7%) |

Data are n, n (%), or median (IQR).

### 3.3 Model performance on the internal cohort

We first evaluated the impact of pretraining on AI-ECG performance for predicting LVEF in the internal test cohort. To simulate low-resource training conditions, we randomly sampled 100, 1,242 (1%), 12,426 (10%), and 124,265 (100%) training instances from the full training set and trained ECG-Fyler and a baseline trained from scratch (“ResNet (Scratch)”) under identical settings.

As shown in Fig. 2a, pretraining substantially improved performance in data-limited regimes. For example, for the LVEF ≤ 50% classification task, pretraining increased AUROC by more than 20 points when only 100 training samples were available. The advantage of ECG-Fyler pretraining was also observed across all LVEF thresholds.

**Fig. 1.**
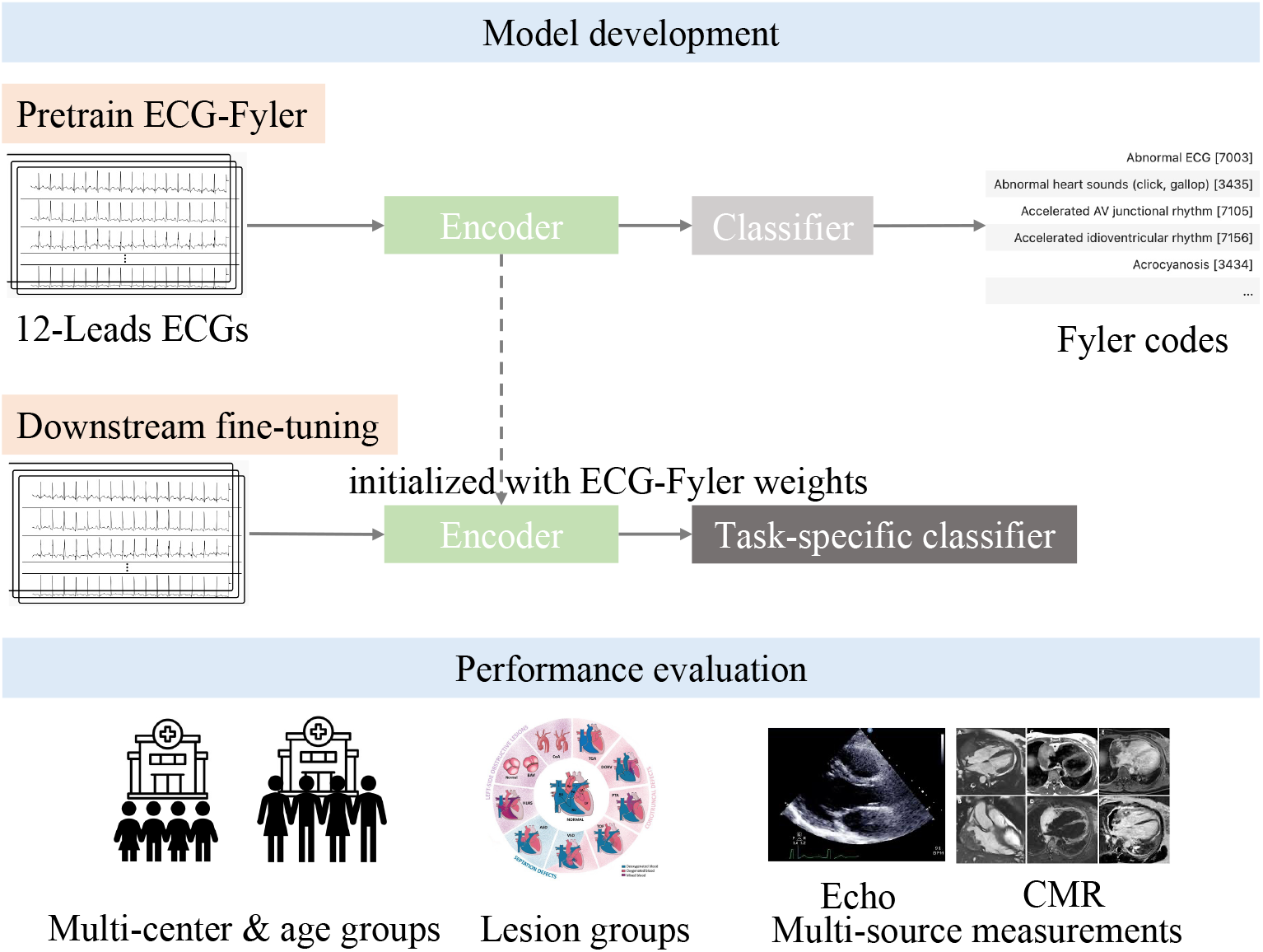
ECG-Fyler development and performance evaluation workflow. **Model development (top):** In the pretraining phase, 12-lead ECGs are used to train ECG-Fyler, an encoder-classifier architecture that predicts Fyler codes. In the downstream fine-tuning phase, the encoder is initialized with ECG-Fyler weights and task-specific classifiers are trained to predict target cardiac phenotypes. **Performance evaluation (bottom):** Models are evaluated across multiple dimensions: multicenter cohorts and age groups, cardiac lesion groups, and multisource measurements including echo and CMR for comprehensive clinical validation.

**Fig. 2.**
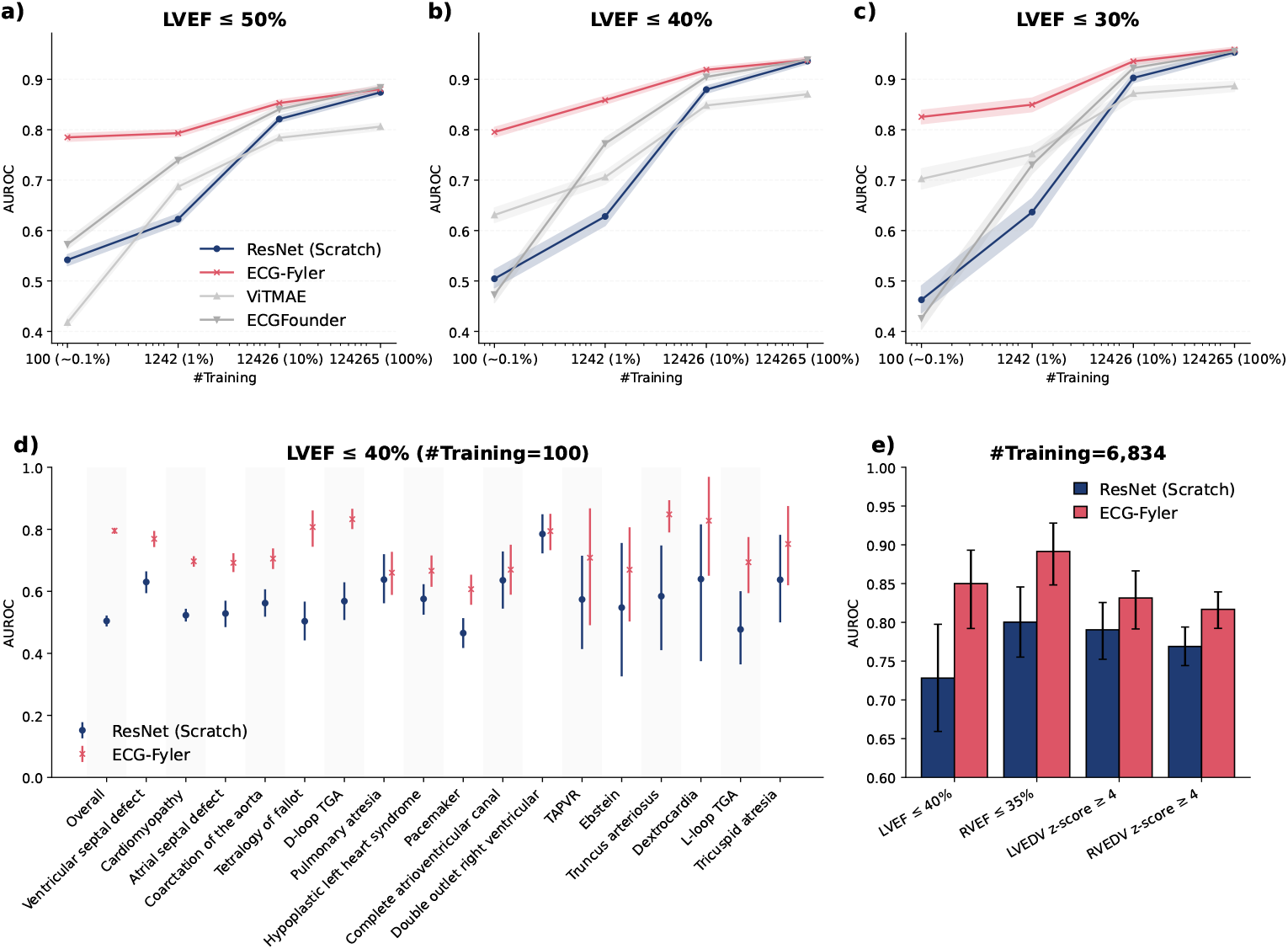
Internal testing of AI-ECG models. **a–c)** Learning curves showing AUROC across randomly sampled training sample sizes (100 ~ [0.1%], 1,242 [1%], 12,426 [10%], 124,265 [100%]) for three LVEF thresholds (≤ 50%, ≤ 40%, ≤ 30%) in ResNet models initialized from scratch (blue), pre-trained weights (ECG-Fyler; red), and baseline approaches (ViTMAE, ECGFounder; gray). **(d)** AUROC for ResNet (Scratch) vs ECG-Fyler across 17 clinical subgroups with LVEF ≤ 40% and fixed training set (n=100). **(e)** Comparative AUROC performance for four CMR-derived measurements: LVEF ≤ 40%, RVEF ≤ 35%, LVEDV z-score ≥ 4, and RVEDV z-score ≥ 4, with full training data (n=6,834). All results show median AUROC with 95% confidence intervals derived from 1,000 bootstrap resamples.

When compared with other foundation models, including the self-supervised ViT-MAE and the general-pretrained ECGFounder, ECG-Fyler consistently achieved higher performance, particularly under limited-resource conditions (100 or 1,242 training samples; 2). For the LVEF ≤ 40% task, ECG-Fyler achieved a median AUROC of 0.80 (95% CI 0.79–0.80) with 100 training samples, compared with 0.50 (0.49–0.52) for ResNet trained from scratch, 0.63 (0.62–0.64) for ViTMAE, and 0.47 (0.46– 0.49) for ECGFounder. At 1% of the training data, the corresponding AUROCs were 0.86 (0.85–0.87), 0.63 (0.61–0.64), 0.71 (0.69–0.72), and 0.77 (0.76–0.78), respectively. These results suggest that supervised pretraining on paediatric ECGs provides advantages over self-supervised pretraining. While ViTMAE was pretrained on the same paediatric cohort using a masked reconstruction objective, it consistently underperformed compared with ECG-Fyler, particularly under low-resource training conditions. Additionally, supervised pretraining on more general annotations in adults, as in ECGFounder, shows limited generalization to paediatric ECGs.

The performance gap gradually narrowed as the training set size increased, with ECG-Fyler and ResNet trained from scratch showing comparable results when the full training set was available. Notably, ECG-Fyler achieved performance comparable to using the entire dataset with substantially fewer training samples. Receiver operating characteristic curves of the same models are shown in Fig. 3.

**Fig. 3.**
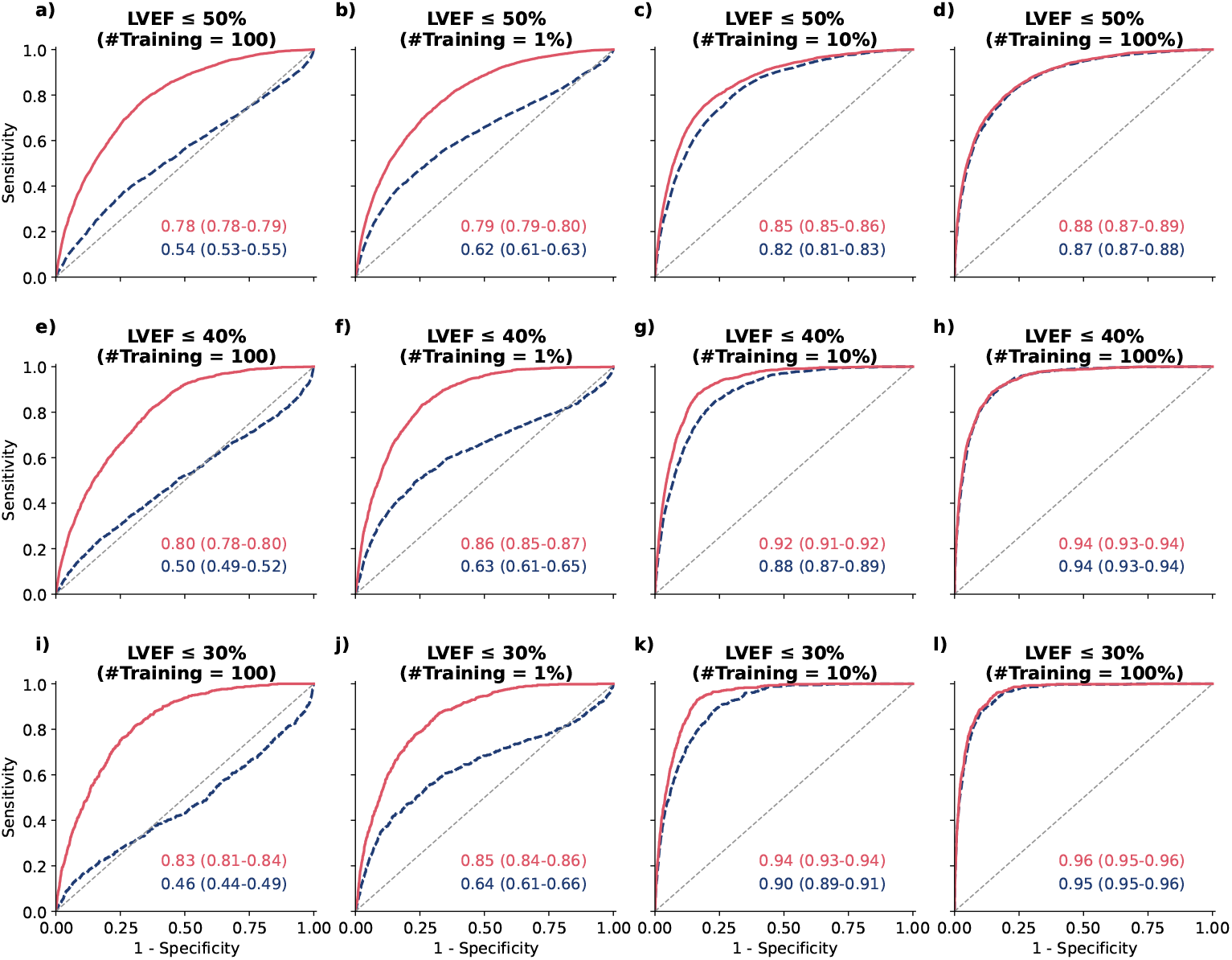
Receiver operating characteristic (ROC) curves for LVEF thresholds across varying training data sizes on the internal test set. **(a–d)** LVEF ≤ 50%, **(e–h)** LVEF ≤ 40%, and **(i–l)** LVEF ≤ 30%, evaluated under different proportions of training data (n=100, 1%, 10%, and 100%). Red solid curves denote models initialized with pre-trained weights, while blue dashed curves denote models trained from scratch. The numerical values shown within each panel correspond to AUROC (with 95% confidence intervals estimated via 1,000 bootstrap resamples), reported separately for the pre-trained (red) and scratch (blue) models.

Because CMR data capture is more intensive, CMR datasets naturally represent low-resource scenarios (6,834 training samples). As a result, ECG-Fyler consistently improved AUROC across all CMR-derived measures compared to task-specific training, including LVEF *≤* 40%, RVEF *≤* 35%, LVEDV z-score ≥ 4, and RVEDV z-score ≥ 4, shown in Fig. 2e.

### 3.4 Subgroup analyses

We further examined model performance across lesion subgroups with markedly different prevalences. For example, only 0.9% of the cohort had tricuspid atresia, highlighting the substantial imbalance across CHD phenotypes (Fig. 2d). Evaluating model performance within these heterogeneous lesion groups therefore provides a more direct assessment of generalization across populations with specific disease classifications.

Fig. 2d shows the effect of pretraining on LVEF ≤ 40% prediction across lesion groups when only 100 training samples were available. Overall, pretraining improved AUROC across all lesion subgroups.

The most pronounced gain was observed in the tetralogy of Fallot group, where the median AUROC increased from 0.50 (95% CI 0.44–0.57) with ResNet trained from scratch to 0.81 (0.74–0.86) with ECG-Fyler. Notably, for patients with pacemakers, the ECG is dominated by pacing artifacts and non-physiologic ventricular activation, which may obscure native conduction and repolarization patterns linked to ventricular function. As a result, LVEF prediction in this cohort represents a greater challenge, but also serves as an important proof-of-concept for the robustness and generalizability of ECG-Fyler. For patients with pacemakers, AUROC likewise improved from 0.47 (0.42–0.51) to 0.61 (0.56–0.65). Given that pacemaker ECGs are dominated by pacing-induced patterns rather than native conduction, these results indicate that pretraining can enhance model robustness specifically in lesion groups with atypical ECG characteristics.

### 3.5 Model performance on the external cohort

We evaluated model performance on an external adult cohort from a different institution, while the pretrained weights were learned exclusively from an internal paediatric cohort with a markedly distinct disease spectrum and ECG distribution (Fig. 4). Despite this substantial domain shift, ECG-Fyler conferred consistent and pronounced gains across all three LVEF thresholds. In the extreme low-data regime (n=100, 0.1%), ECG-Fyler outperformed models trained from scratch (ΔAUROC = 0.13, 0.13, and 0.15 for LVEF ≤ 50%, ≤ 40%, and ≤30%, respectively), indicating strong cross-domain generalization of learned representations. As training data increased, the performance gap narrowed, with both approaches converging at full data. However, ECG-Fyler remained consistently equal to or slightly superior, suggesting that paediatric-derived representations capture physiologically meaningful features that generalize across age groups, institutions, and acquisition protocols.

**Fig. 4.**
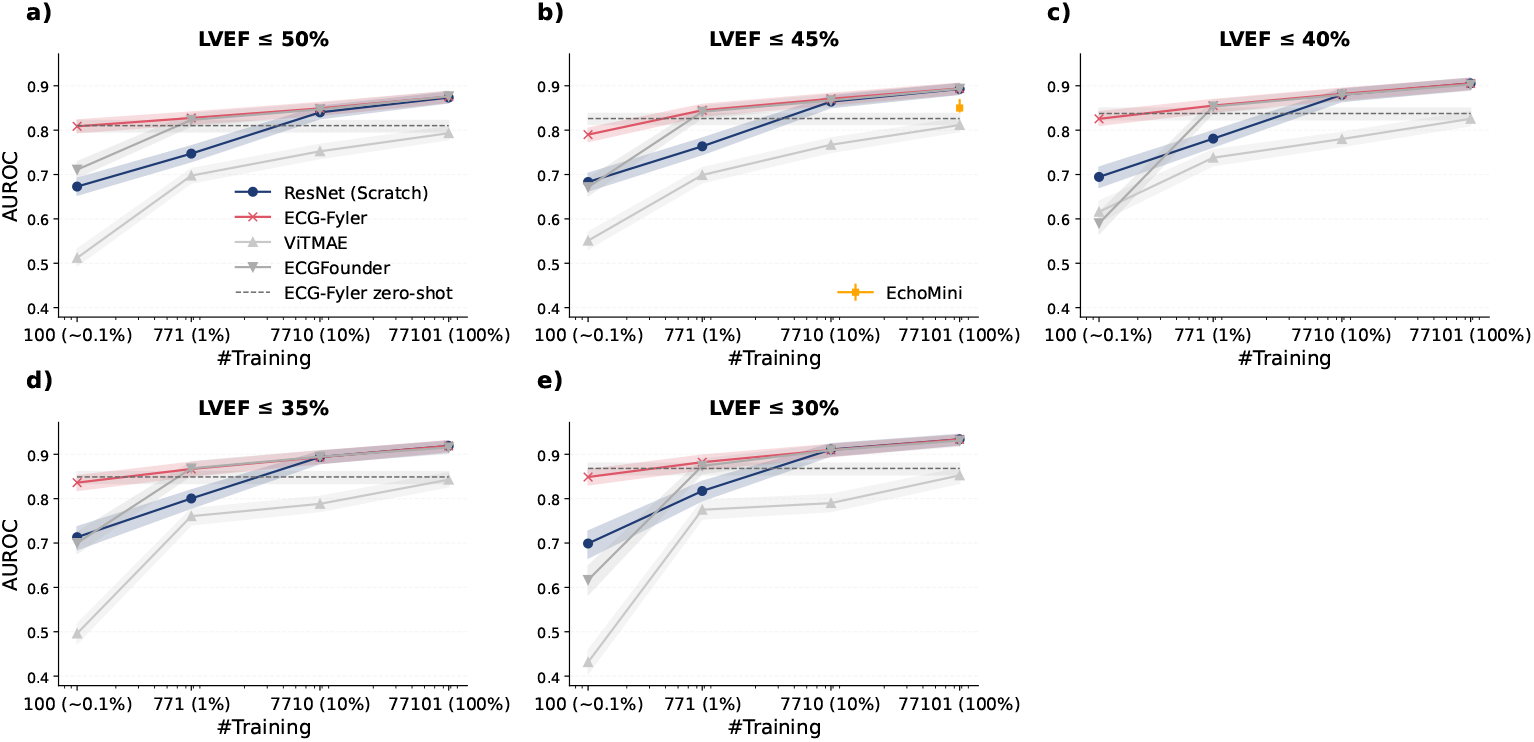
External testing of the AI-ECG models. **(a–e)** Learning curves showing median AUROC across randomly sampled training sample sizes (100 ~ [0.1%], 771 [1%], 7,710 [10%], 77,101 [100%]) for five LVEF thresholds in ResNet models initialized from scratch (blue), pre-trained weights (ECG-Fyler; red), and baseline approaches (ViTMAE, ECGFounder; gray). The dashed line represents ECG-Fyler zero-shot performance, which evaluates the model trained on the internal cohort directly on the external cohort without any fine-tuning. EchoMini results are available only for LVEF ≤ 45% from the original paper ^[15,25]^ with different bootstrap resampling. All results show median AUROC with 95% confidence intervals derived from 1,000 bootstrap resamples.

Compared with ECGFounder, which was supervised-pretrained on 10.7 million adult ECGs from the Harvard-Emory database (approximately 14x larger than our paediatric pretraining dataset), ECG-Fyler achieved comparable performance across tasks. Notably, under extremely low-resource conditions (100 training samples), ECG-Fyler significantly outperformed ECGFounder (AUROC: 0.81, 95% CI: [0.80— 0.82] vs. 0.72 [0.70-–0.73]). This cross-cohort transfer highlights the effectiveness of paediatric-supervised pretraining for generalization to adult populations and supports data-efficient deployment.

In direct external validation, ECG-Fyler achieved strong discrimination on the external cohort, with median AUROC values of 0.81, 0.84, and 0.87 for identifying LVEF ≤ 50%, ≤ 40%, and ≤ 30%, respectively. Notably, this “zero-shot” performance was comparable to, and in some cases even exceeded, that obtained when fine-tuning with a very limited number of external labels (n = 100).

### 3.6 Visual Analysis of ECG Representations Across Cohorts

We investigated the structure of learned ECG representations across cohorts using Uniform Manifold Approximation and Projection (UMAP) dimensionality reduction. In the joint embedding space (Fig. 5a), adult CUIMC embeddings largely occupied regions already populated by BCH embeddings, with substantial overlap across cohorts. A similar pattern was observed when restricting to ECGs with reduced LVEF (LVEF ≤50%; Fig. 5b). Consistent with these representation-level observations, ECG-Fyler generalized to the external CUIMC cohort without additional adaptation, achieving AUROC *>* 0.8 for LVEF prediction across evaluated thresholds (Fig. 4). These results suggest that the learned representations capture robust physiological structure that is transferable across age distributions and institutional settings.

**Fig. 5.**
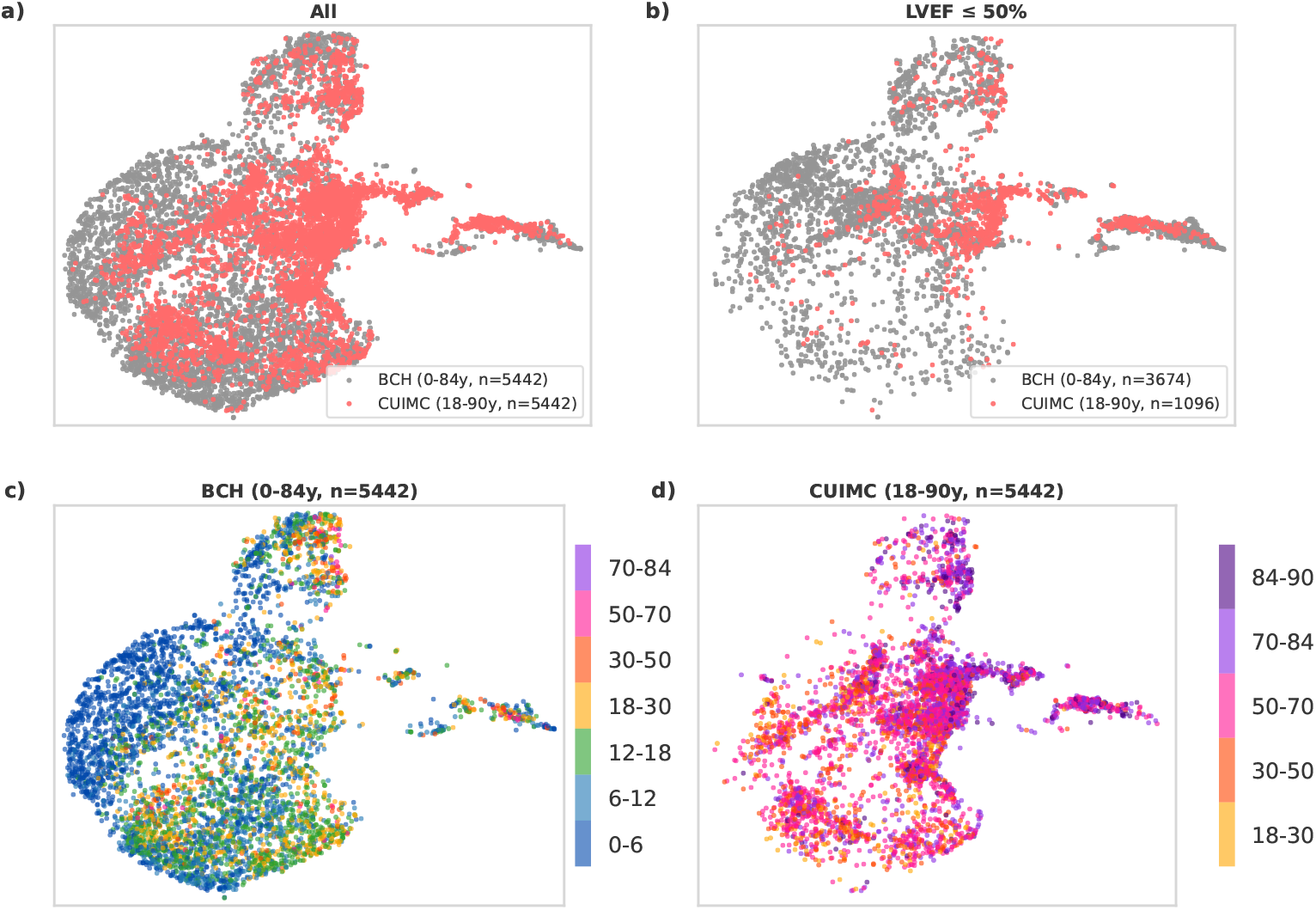
UMAP visualization of ECG embeddings from internal and external cohorts. **(a)** All embeddings from BCH (0–84 years, randomly subsampled to n=5,442) and CUIMC (18–90 years, n=5,442). **(b)** Embeddings restricted to recordings with reduced LVEF (≤ 50%). **(c–d)** BCH- and CUIMC-only embeddings colored by age bins; color bars indicate the corresponding age-group bins.

Age-stratified cohort-specific views showed localized enrichment of younger ages in BCH (Fig. 5c), with the 0-6 years group concentrated in a visually distinct region relative to older paediatric bins, whereas adult age bins in CUIMC were broadly intermixed across the embedding space (Fig. 5d).

## 4 Discussion

Our results show that ECG-Fyler improves cardiac function assessment compared to existing AI-ECG approaches, particularly when data resources are scarce. ECG-Fyler consistently outperformed or matched models trained from scratch, as well as prior adult-derived ECG foundation models. The results suggest that Fyler-code based pretraining, as well as “all-ages” pretraining, improve the data efficiency and transferability of AI-ECG models.

These gains are especially relevant in CHD, where anatomic heterogeneity, lifelong surveillance, and limited labels within individual lesion subgroups complicate model development. ECG-Fyler’s pretraining on a predominantly paediatric, lifespan-spanning cohort annotated with Fyler codes improved generalization across rare lesions and pacemaker-associated rhythms, suggesting that transferable ECG representations can mitigate data scarcity in uncommon anatomies.

Compared with self-supervised pretraining (ViTMAE), supervised pretraining (i.e., ECG-Fyler) achieved higher performance across settings, consistent with the benefit of task-relevant supervision when labels are available at scale. ECG-Fyler also generalized under age and institutional shift, whereas adult-biased pretraining transferred less effectively to paediatric dominant cohorts, with the largest gap in the lowest-resource regimes.

UMAP analyses were consistent with this cross-age transfer. ECG-Fyler embeddings of adult CUIMC ECGs aligned with the global low-dimensional structure learned from BCH ECGs, including within the subgroup with LVEF ≤ 50%. Together with the external classification results, this pattern suggests that all-ages cohorts can help models learn ECG features that remain useful across heterogeneous populations.

From a clinical perspective, these improvements support more reliable ECG-based screening and longitudinal monitoring of ventricular function, including RV dysfunction and ventricular dilation, for which access to advanced imaging (CMR) is often limited. Better discrimination in low-data settings could improve triage by reducing unnecessary imaging while prioritizing patients who warrant echo or CMR imaging, particularly in settings with constrained resources or limited subspecialty access. There is large potential for low-cost, opportunistic screening via ECGs in low-resource settings to help direct more resources to patients with abnormal cardiac function that would normally be missed.

### 4.1 Limitations

Several limitations warrant consideration. First, low-resource settings were simulated by randomly subsampling the training set rather than prospectively evaluating sites with limited labeled data; each site has unique characteristics, care protocols, and patient demographics that may affect model performance. Second, most experiments focused on ventricular function endpoints, particularly LVEF. Broader evaluation across additional ECG tasks and real-world low-label settings will be important to define the full scope of benefit from supervised pretraining on an all age cohort.

## Data Availability

Deidentified participant data collected at Boston Children's Hospital, including the corresponding data dictionary, are not planned for public release because of patient privacy and institutional restrictions. Restricted access requests may be considered from the time of publication for researchers with a methodologically sound proposal by contacting the corresponding authors, subject to institutional review, ethical approval, and a data use agreement. No study protocol, statistical analysis plan, or informed consent form will be made publicly available.

https://physionet.org/content/echonext/1.1.0

https://github.com/cavalab/fyler_code_fm

## Data sharing

Deidentified participant data collected at Boston Children’s Hospital, including the corresponding data dictionary, are not planned for public release because of patient privacy and institutional restrictions. Restricted access requests may be considered from the time of publication for researchers with a methodologically sound proposal by contacting the corresponding authors, subject to institutional review, ethical approval, and a data use agreement. No study protocol, statistical analysis plan, or informed consent form will be made publicly available. The external EchoNext cohort used for external validation is publicly available from the time of publication at https://physionet.org/content/echonext/1.1.0/. Use of Boston Children’s Hospital data for this study was approved by the Boston Children’s Hospital institutional review board. The model weights and code used for data preprocessing, model training, and evaluation is publicly available from the time of publication at https://github.com/cavalab/fyler_code_fm.

## Contributors

WGL and TM conceptualised the project and obtained funding and resources. YY designed the methodology, conducted the primary experiments and formal analysis, and prepared the first draft of the manuscript. JM curated the data and constructed the dataset. LP conducted the baseline comparator model experiments. WGL and TM supervised the study and provided methodological guidance. YY, JM, LP, WGL, and TM reviewed and edited the manuscript and provided critical feedback. YY, TM and WGL had direct access to and verified the underlying data reported in the manuscript. All authors had final responsibility for the decision to submit for publication.

## Acknowledgments

Research reported in this publication was supported by the National Library of Medicine of the National Institutes of Health under Award Number R01LM012973. This work was partially supported by the Kostin Innovation Fund at Boston Children’s Hospital.

