## Supplementary Material for "An ECG foundation model for generalizable cardiac function prediction across the lifespan"

### Web Extra Material

**Supplementary Tables**

**Table S1:** Fyler code distribution in the pretraining cohort (Part 1). Fyler-coded electrocardiogram (ECG) diagnostic categories are listed with their corresponding identifiers and prevalence, reported as counts and percentages of total recordings. Codes are ordered by decreasing frequency.

| | |
|---|---|
| Normal ECG | 332,708 (42.538%) |
| Incomplete right bundle branch block (RSR') | 101,333 (12.956%) |
| ST-T wave change, non-specific | 80,560 (10.300%) |
| Right bundle branch block (complete) | 67,800 (8.669%) |
| Right ventricular hypertrophy (ECG) | 60,619 (7.750%) |
| Axis deviation, right (ECG) | 59,606 (7.621%) |
| Axis deviation, superior (ECG) | 36,500 (4.667%) |
| QTc prolonged | 35,974 (4.599%) |
| Sinus tachycardia | 35,971 (4.599%) |
| Sinus arrhythmia | 34,005 (4.348%) |
| Axis deviation, left (ECG) | 28,149 (3.599%) |
| Intraventricular conduction delay | 28,009 (3.581%) |
| Right atrial enlargement (ECG) | 25,867 (3.307%) |
| Sinus bradycardia | 25,797 (3.298%) |
| Probable right ventricular hypertrophy (ECG) | 23,547 (3.011%) |
| Diminished LV (or lateral) forces (ECG) | 23,361 (2.987%) |
| First degree atrioventricular block | 22,922 (2.931%) |
| Left ventricular hypertrophy (ECG) | 22,560 (2.884%) |
| Dual chamber pacing | 18,059 (2.309%) |
| Ectopic atrial rhythm | 18,029 (2.305%) |
| T-wave inversion | 16,341 (2.089%) |
| Probable left ventricular hypertrophy (ECG) | 13,250 (1.694%) |
| Premature ventricular beats (unifocal) | 11,615 (1.485%) |
| Left atrial enlargement (ECG) | 11,383 (1.455%) |
| Counterclockwise rotation (ECG) | 11,201 (1.432%) |
| Atrial abnormality (ECG) | 10,891 (1.392%) |
| Atrial pacing | 10,125 (1.295%) |
| Ventricular hypertrophy, non-specific (ECG) | 9,097 (1.163%) |
| Premature atrial beats | 7,931 (1.014%) |
| Early repolarization | 7,510 (0.960%) |
| Wolff Parkinson White syndrome | 7,374 (0.943%) |
| Strain pattern - ST-T, right | 6,961 (0.890%) |
| Probably normal ECG variant | 6,735 (0.861%) |
| Diminished RV (or anterior) forces (ECG) | 6,450 (0.825%) |
| Biatrial enlargement (ECG) | 6,083 (0.778%) |
| Junctional escape beats | 6,060 (0.775%) |
| Ventricular pacing | 5,876 (0.751%) |
| Technically inadequate study | 5,718 (0.731%) |
| Strain pattern - ST-T, left | 5,160 (0.660%) |
| Low voltage (ECG) | 4,553 (0.582%) |
| Left bundle branch block | 4,111 (0.526%) |
| Atrial fibrillation | 3,331 (0.426%) |
| Atrial flutter | 2,982 (0.381%) |
| Short PR interval | 2,915 (0.373%) |
| Accelerated AV junctional rhythm | 2,860 (0.366%) |
| Dextrocardia (ECG) | 2,835 (0.362%) |
| Left anterior hemiblock | 2,506 (0.320%) |
| Complete heart block | 2,169 (0.277%) |

**Table S2:** Fyler code distribution in the pretraining cohort (Part 2).

| | |
|---|---|
| ST-T wave abnormalities, injury or ischemia | 2,020 (0.258%) |
| Ventricular inversion (ECG) | 2,012 (0.257%) |
| ST-T wave abnormality, pericardial disease | 1,948 (0.249%) |
| Supraventricular tachycardia narrow QRS | 1,606 (0.205%) |
| Automatic ectopic atrial tachycardia | 1,579 (0.202%) |
| Wandering pacemaker | 1,445 (0.185%) |
| Potentially serious ECG, cardiology evaluation required | 1,258 (0.161%) |
| Left ventricular conduction delay | 1,117 (0.143%) |
| Wenckebach (Mobitz type 1), second degree atrioventricular block | 979 (0.125%) |
| Miscellaneous ECG codes | 754 (0.096%) |
| Pacemaker malfunction | 631 (0.081%) |
| Supraventricular tachycardia wide QRS | 573 (0.073%) |
| High grade atrioventricular block | 547 (0.070%) |
| Probably abnormal ECG for age, clinical correlation advised | 542 (0.069%) |
| Situs inversus (ECG) | 502 (0.064%) |
| Atrioventricular dissociation | 484 (0.062%) |
| Myocardial infarction ECG pattern | 479 (0.061%) |
| Pathologic Q waves | 463 (0.059%) |
| ECG Hypokalemia | 439 (0.056%) |
| Junctional premature beats | 420 (0.054%) |
| Accelerated idioventricular rhythm | 416 (0.053%) |
| Couplets ventricular origin | 404 (0.052%) |
| Sinus arrest or sino-atrial block or pause | 399 (0.051%) |
| Mobitz type 2 | 362 (0.046%) |
| Ventricular escape beat | 353 (0.045%) |
| Ventricular tachycardia sustained | 314 (0.040%) |
| Ventricular tachycardia non sustained | 305 (0.039%) |
| Automatic junctional tachycardia post op | 267 (0.034%) |
| ECG Hypocalcemia | 259 (0.033%) |
| First degree Atrioventricular block | 203 (0.026%) |
| Biventricular hypertrophy (ECG) | 159 (0.020%) |
| ECG Drug effect | 158 (0.020%) |
| Automatic junctional tachycardia | 153 (0.020%) |
| ECG Hyperkalemia | 110 (0.014%) |
| Pacemaker | 105 (0.013%) |
| ECG Digoxin effect | 70 (0.009%) |
| Multifocal atrial tachycardia | 67 (0.009%) |
| Mahaim fiber | 65 (0.008%) |
| Left posterior hemiblock | 59 (0.008%) |
| Junctional rhythm nodal | 39 (0.005%) |
| Complete heart block (Acquired) | 32 (0.004%) |
| Pulmonary vein abnormality | 22 (0.003%) |
| Infarct, anteroseptal | 16 (0.002%) |
| Supraventricular tachycardia | 16 (0.002%) |
| Supraventriculr tachycardia wide QRS | 10 (0.001%) |

**Supplementary Figures**

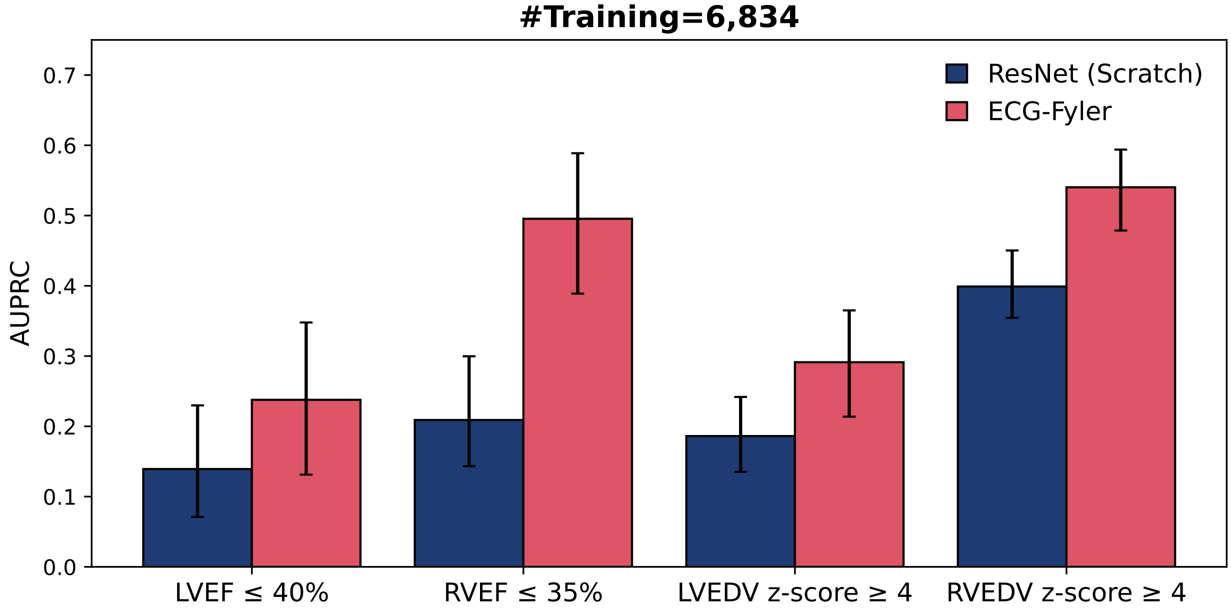

**Figure S1:** Area under the precision-recall curve (AUPRC) performance for four cardiac magnetic resonance (CMR)-derived measurements: left ventricular ejection fraction (LVEF) $\leq$ 40%, right ventricular ejection fraction (RVEF) $\leq$ 35%, left ventricular end-diastolic volume (LVEDV) z-score $\geq$ 4, and right ventricular end-diastolic volume (RVEDV) z-score $\geq$ 4, with full training data (n=6,834). All results show median AUPRC with 95% confidence intervals derived from 1,000 bootstrap resamples.

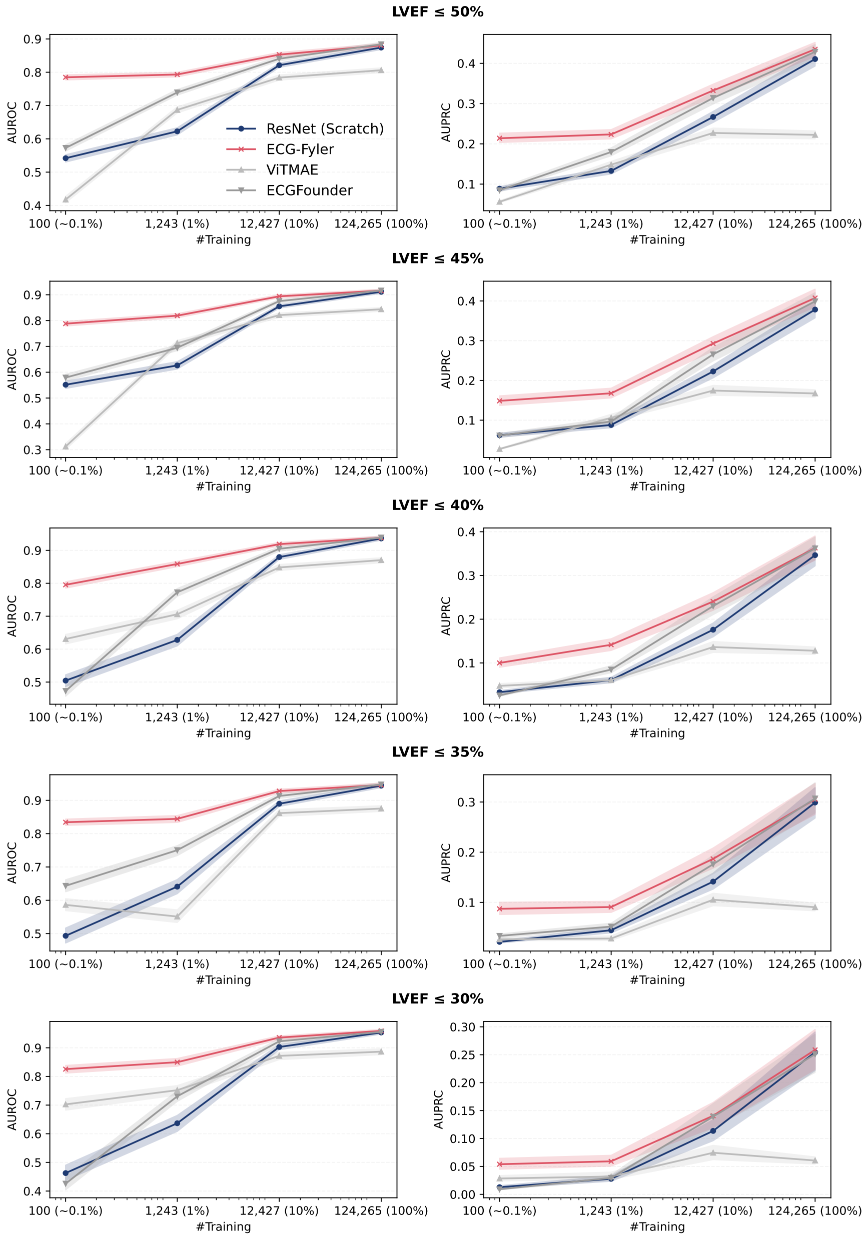

**Figure S2:** Learning curves showing AUROC and AUPRC across randomly sampled training sample sizes (100 [∼0.1%], 1,242 [1%], 12,426 [10%], 124,265 [100%]) for five LVEF thresholds ($\leq$ 50%, $\leq$ 45%, $\leq$ 40%, $\leq$ 35%, $\leq$ 30%) in ResNet models initialized from scratch (blue), pretrained weights (ECG-Fyler; red), and baseline approaches (Vision Transformer Masked AutoEncoder [ViTMAE], ECGFounder; gray).

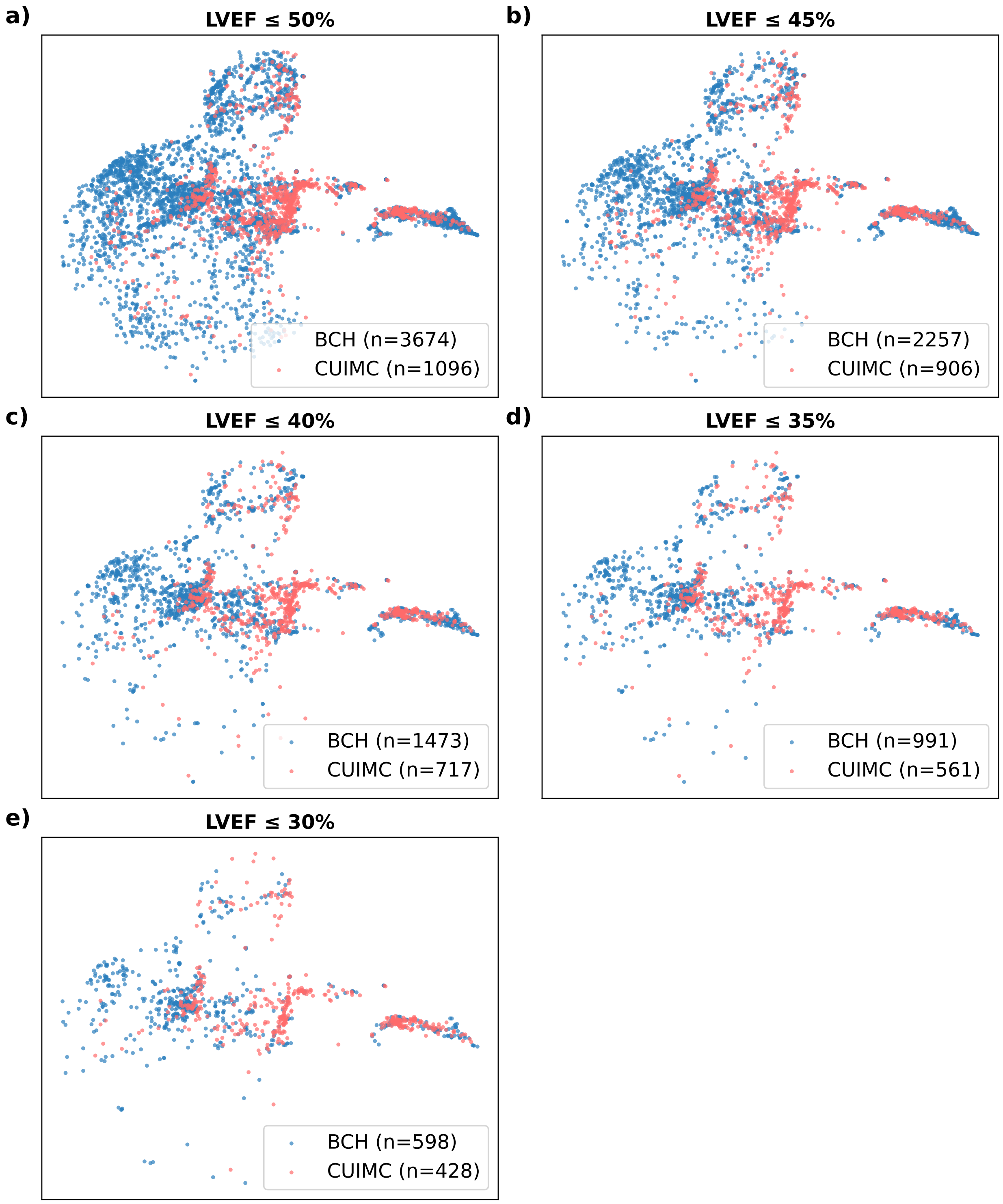

**Figure S3:** Uniform Manifold Approximation and Projection (UMAP) visualization of ECG embeddings from internal and external cohorts. Embeddings from the internal Boston Children's Hospital (BCH) cohort (blue) and the external Columbia University Irving Medical Center (CUIMC) cohort (red) are shown. Each panel corresponds to a different LVEF threshold and displays only positive cases.
